# The Mezurio smartphone application: Evaluating the feasibility of frequent digital cognitive assessment in the PREVENT dementia study

**DOI:** 10.1101/19005124

**Authors:** Claire Lancaster, Jasmine Blane, Amy Chinner, Leona Wolters, Ivan Koychev, Chris Hinds

## Abstract

**Background:** Smartphones may significantly contribute to the detection of early cognitive decline at scale by enabling remote, frequent, sensitive, economic assessment. Several prior studies have sustained engagement with participants remotely over a period of a week; extending this to a period of a month would clearly give greater opportunity for measurement. However, as such study durations are increased, so too is the need to understand how participant burden and scientific value might be optimally balanced.

**Objectives:** We explore the ‘little but often’ approach to assessment employed by the Mezurio app, interacting with participants every day for over a month. We aim to understand whether this extended remote study duration is feasible, and which factors might promote sustained participant engagement over such study durations.

**Methods:** Thirty-five adults (aged 40-59 years) with no diagnosis of cognitive impairment were prompted to interact with the Mezurio smartphone app platform for up to 36 days, completing short, daily episodic memory tasks in addition to optional executive function and language tests. A subset (*n*=20) completed semi-structured interviews focused on their experience using the app.

**Results:** Average compliance with the schedule of learning for subsequent memory test was 80%, with 88% of participants still actively engaged by the final task. Thematic analysis of participants’ experiences highlighted schedule flexibility, a clear user-interface, and performance feedback as important considerations for engagement with remote digital assessment.

**Conclusions:** Despite the extended study duration, participants demonstrated high compliance with the tasks scheduled and were extremely positive about their experiences. Long durations of remote digital interaction are therefore definitely feasible, but only when careful attention is paid to the design of the users’ experience.

## Introduction

Advances in smartphone-based cognitive assessment will significantly benefit the development of clinical interventions for Alzheimer’s disease (AD); facilitating both the detection of covert pathology at an early, preclinical stage, and more exact monitoring of disease progression and therapeutic response (Gold et al., 2018). Inbuilt smartphone sensors can record sensitive trial-level outcomes, plus performance metadata, which may assist in the detection of subtle cognitive impairment. Furthermore, mobile technologies uniquely enable frequent, longitudinal assessment, with significantly lower participant burden and administration cost associated with remote, digital data collection than repeat in-clinic tests (Areán, Ly, & Andersson, 2016; Resnick & Lathan, 2016). By facilitating greater breadth of behavioural assessment, smartphones increase the reliability of cognitive profiling, reducing the impact of within-participant variability, for example in association with daily stressors, mood and sleep (e.g. Brose, Schmiedek, Lövdén, & Lindenberger, 2012; Hess, Popham, Emery & Elliott, 2012; Schmidt, Collette, Cajochen, & Peigneux, 2007). In addition, frequent measurement allows acute longitudinal change in cognitive function to be accurately assessed (Koo & Vizer, 2019). Although digital technology promises clear benefits for dementia research, usability is the greatest challenge to these tools being widely adopted (Gold et al., 2018); this research directly addresses this barrier.

The feasibility of remote, digital cognitive assessment in older adults is commonly assessed according to study compliance (Klimova, 2017; Koo & Vizer, 2019; Onoda & Yamaguchi, 2014; Ruggeri, Maguire, Andrews, Martin, & Menon, 2016). Emphasis has been placed on deploying short, frequent assessments, with adherence to a schedule of self-reported function and active behavioural tasks in the range of 72% - 82% when older adults with no objective cognitive impairment were prompted to interact 5 times a day for a week (Allard et al., 2014; Schweitzer et al., 2017). Further support for the acceptability of high frequency ‘micro-interactions’ (<1 minute) is evidenced by 77% compliance in older adults prompted via smartphone notification to complete four assessments at random intervals across each day for a week (Lange & Süß, 2014). Tests of longitudinal adherence across six months in individuals at increased familial risk of dementia reported participation attrition (8.1%) at longer study durations (Jongstra et al., 2017), emphasising the importance of strategically promoting long-term participant engagement to maximise data quality.

Despite the significance of participant compliance for the value of research outcomes, there has been limited evaluation of which factors promote sustained engagement with repeated, digital cognitive assessment. A focus-group discussion between young and older adults highlighted participant autonomy in scheduling, positive feedback, and specific instructions on how to approach each cognitive task as important factors for engagement (Jenkins, Lindsay, Eslambolchilar, Thornton, & Tales, 2016). More broadly, for sustaining long-term engagement (3-17 months) with an online health platform, older adults highlighted personalised reminders from the platform, incorporating the tool into their daily routine, and observed variation or progress within the platform as important (Van Middelaar et al., 2018).

The current study tested the feasibility of the Mezurio smartphone app (https://mezur.io); specifically, the utility of this tool for significantly longer-term, high-frequency assessment (≤ 36 days) than the seven-day assessment periods explored previously (Allard et al., 2014; Lange & Süß, 2014; Schweitzer et al., 2017), and which factors contribute to successful participant engagement. Mezurio contains a collection of novel behavioural tasks measuring long-term episodic memory, language and executive function through a range of input modalities including voice, movement and touch; each targeted at the detection of preclinical AD. These tasks follow a comparable ‘little and often’ approach to remote assessment as tested previously (Allard et al., 2014; Schweitzer et al., 2017; Sliwinski et al., 2018), with emphasis placed on providing participants autonomy to schedule tasks according to their daily routine. In addition, the developers have worked closely with lay older adults (including adults with mild cognitive impairment) to create a research experience anticipated to be both clear and engaging.

Here, mid-age participants were prompted to interact with an extended schedule of daily cognitive assessments within the Mezurio app, with feasibility objectively tested according to study compliance and participant attrition. Semi-structured interviews, focused on themes of approachability, acceptability and engagement, were used to evaluate which factors contribute to participants willingness to engage in digital cognitive assessments for longer durations. This proof of concept is essential for establishing app utility ahead of inclusion of Mezurio in larger clinical trials, plus improving the use of digital tools in health research more broadly.

## Methods

### Participants

Descriptive data for 35 adult volunteers (97% Caucasian, aged 40-59 years) are shown in Table 1, including for the subset of participants (*n*=20) who completed the semi-structured interview following the period of smartphone assessment. There were no significant group differences in the characteristics of participants providing qualitative feedback and those not completing the interview (independent-groups *t-*tests: age (*p*=.845), years of educations (*p*=.683); X_2_ tests: gender (*p*=.359), immediate family history of dementia (*p*=.192). Participants were recruited from the Oxford study-site of the PREVENT dementia programme (Ritchie & Ritchie, 2012), which excluded individuals with diagnosable dementia. This prospective research study is investigating the interactions between risk factors for dementia and traditional biomarkers in mid-life. Although family history of dementia was not an inclusion criterion, participants appeared to self-select for this characteristic: a high proportion (66%) reported a first-degree relative with dementia (43% AD). The current study was ethically approved (University of Oxford Medical Sciences Inter-Divisional Research Ethics Committee: R48717/RE001) and compliant with the Helsinki Declaration of 1975. Written consent was required upon entry to the study.

**Table 1.** Demographic characteristics of participants

| **Demographics** | **Total sample**<br>**(*n*=35)** | **Interview sub-group**<br>**(*n*=20)** |
| --- | --- | --- |
| **Age (years)** | 52.57 (5.10) | 52.20 (5.15) |
| **Gender (% Female)** | 74 | 80 |
| **Years of Education** | 15.49 (2.74) | 15.30 (3.16) |
| **Family History (%)** | 66 | 75 |

### Mezurio Smartphone Assessments

Participants were asked to complete a selection of freely available cognitive tasks within the Mezurio smartphone app platform (https://mezur.io), each intended to take less than five minutes to complete, with ongoing longitudinal follow-up scheduled at 6- and 12-months. Two of these tasks (Tilt Task, Story Time) were only introduced once a subset of participants (*n*=23) had completed their baseline month of assessment, with subsequent recruits (*n*=12) being asked to complete the full selection of tasks. At month 6, the first 23 participants were offered the opportunity to switch to this more comprehensive version of the app, with 19 opting to make this change.

### Cognitive measures

All participants in the current study completed Gallery Game (Lancaster et al., 2019), composed of regular cross-modal paired-associates learning tasks with subsequent tests of recognition and recall following ecologically relevant delays (for the current mid-age population specified as 1, 2, 4, 6, 8, 10, or 13 days). Within each learning activity, participants were asked to encode distinct pairings of object photo-stimuli and touchscreen ‘swipe’ directions (left, right or up), with the number of object-direction associations progressively increasing alongside iterative checks of immediate recall for pairings until the learning criterion was achieved (for more detail see Lancaster et al., 2019). Positive feedback (a gold star animation) was presented following each learning iteration if immediate recall was 100% correct. Incorrect responses were indicated by the object-direction association being cued so the participant could repeat the learning trial, followed by an immediate second test of recall.

Participants did not receive explicit feedback on recognition or recall test performance. Excluding practice trials, participants were prompted to complete up to 22 learning tasks, plus the associated recognition and recall tests for encoded stimuli. Participants interacted with Gallery Game once a day for up to 29 days.

In addition, participants completing the extended version of Mezurio were prompted to complete Story Time and Tilt Task. Story Time provides a test of ‘connected’ language, with participants asked to narrate a short comic strip aloud whilst the device microphone was recording and then repeat the story from memory immediately and following an ∼ 24-hour delay. No feedback on performance is provided by Mezurio. Tilt task, a measure of executive function, relies on device movement sensors. Participants were asked to tilt their phone in order to move the central ‘cursor’ towards the next target in sequence, with executive challenge increasing with successive levels. Inaccurate responses were registered by the central cursor rebounding off the non-target lure and participants losing one of their 3 lives. Positive feedback on performance is symbolized by the collection of trophies as participant’s move through the levels. Note, if a participant does not reach the end of the level they cannot move onto the next. Figure 1 provides a visual representation of all three tasks.

**Figure 1.**
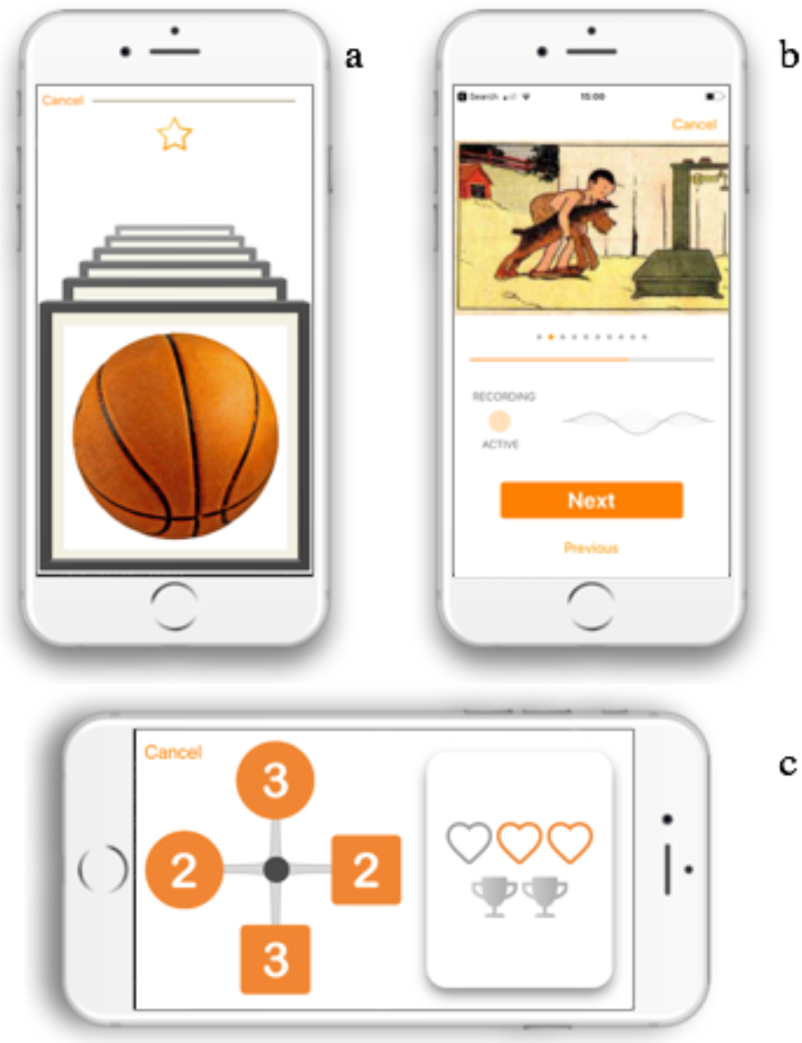
Cognitive tasks deployed within Mezurio: a) Gallery Game, b) Story Time, and c) Tilt Task.

### Procedure

Participants were invited to download the Mezurio app onto their personal smartphone (Apple or Android) or a loaned Android device, with written instructions for download and a unique authentication ID provided by the research team. Mezurio asked participants to complete daily tasks within the app for up to 36 days. Specifically, the Gallery Game-only version of Mezurio asked participants to interact once a day, however, the schedule for the extended version varied by including: 1) additional Story Time assessments alongside Gallery Game for 7 consecutive days; 2) a final week of Tilt Task presented three times per day (e.g. after breakfast, lunch and dinner). Note, 6- and 12-month follow-up assessment within Mezurio is ongoing, with the current study providing a first look at method feasibility after completion of all baseline assessments.

The app encouraged participants to complete the cognitive tasks at the same time each day via a phone-based notification, with a second reminder 15-minutes later if the task was not initiated. Participants had 16 hours to complete each activity before the task ‘expired’, with the exception of Tilt Task which must be completed within 2 hours. Participants chose the time of their scheduled tasks when first opening Mezurio; these notification times could be changed at any time from within the home screen of the app.

### Semi-structured interviews

Participants were invited to provide feedback on their experiences using the Mezurio smartphone app, with the first 20 volunteers selected to complete a semi-structured interview (duration: ∼30 minutes). At the time of interview all participants had completed their baseline period of assessments, with 13 participants having completed or in the process of completing their 6-month follow-up. Four participants had experience limited to the Gallery Game episodic memory task only, with a further 3 participants unable to complete Tilt Task as their smartphone was not compatible. The schedule of interview questions (Appendix) included a mixture of closed-answer questions requiring a score out of 10 and open-ended questions, with prompts tailored to each participant’s responses. The presentation of questions broadly mapped to the order in the schedule, with revisions according to individual responses. Interviews were audio recorded and transcribed verbatim.

### Data analysis

As all participants were prompted to complete a schedule of daily Gallery Game tasks during the baseline assessment period, compliance was selectively considered for this programme of activity, excluding the secondary Tilt Task and Story Time measures. Here, compliance is calculated as the proportion of scheduled object-direction pairings attempted during learning. The association between participant characteristics and learning compliance (reverse-score square root transformation to account for negative skew) was screened, using between-groups t-tests for categorical (immediate family history, gender) and simple linear regression for continuous (age, years of education) variables. In addition, attrition to study participation was proxied as how far through the schedule of Gallery Game learning tasks were participants’ last active. Note, the analysis of compliance emphasises learning of object-direction pairings as opposed to recognition and recall as memory tests were only presented by Mezurio if the associated learning activity had been successfully completed, hence these measures were not independent.

Interviews were subject to thematic analysis (Braun & Clarke, 2006), with the lead author (CL) taking a deductive approach to analysing the transcripts focused on the following research themes: 1) approachability of Mezurio app, including a more general consideration of smartphone technology, 2) acceptability of the research ask and 3) engagement with the cognitive tasks. Additional themes emerging from the data were considered, but are not considered here as factors contributing to the feasibility of smartphone-based assessment. The lead author (CL) read each transcript iteratively to extract words and phrases representing key themes of participant experience.

## Results

### Compliance and attrition

Average compliance to the schedule of Gallery learning tasks was 77.85±15.88%. One outlier (4.55%) only completed the first learning task, later withdrawing from the study due to the time commitment. Excluding this participant, average compliance is 80.00±9.60%. An immediate family history of dementia (*M*=79.13 ± 9.33) was associated with poorer compliance to the study schedule in comparison to those with no family history (*M*=81.79 ± 10.38), *t*(43.33)=17.25, *p*<.001); as was being female (*M*=79.30 ± 9.38) as opposed to male (*M*=82.30 ± 10.62, *t*(41.67)=12.41, *p*<.001). These associations must be considered with caution, however, due to limited group sizes. There was no significant association between age *(p*=.096) and years of education *(p*=.741) with compliance to the schedule of Gallery Game learning tasks. Participants completed an average of 67.26 ± 20.61% recognition tests and 66.63 ± 20.59% recall tests (see Figure 2). Attrition across the baseline period of Gallery learning tasks was limited, with 88% of participants completing the final scheduled learning task. The mean proportion of the schedule completed at the point of final learning was 98.15±5.70%, range 75 – 100%.

**Figure 2.**
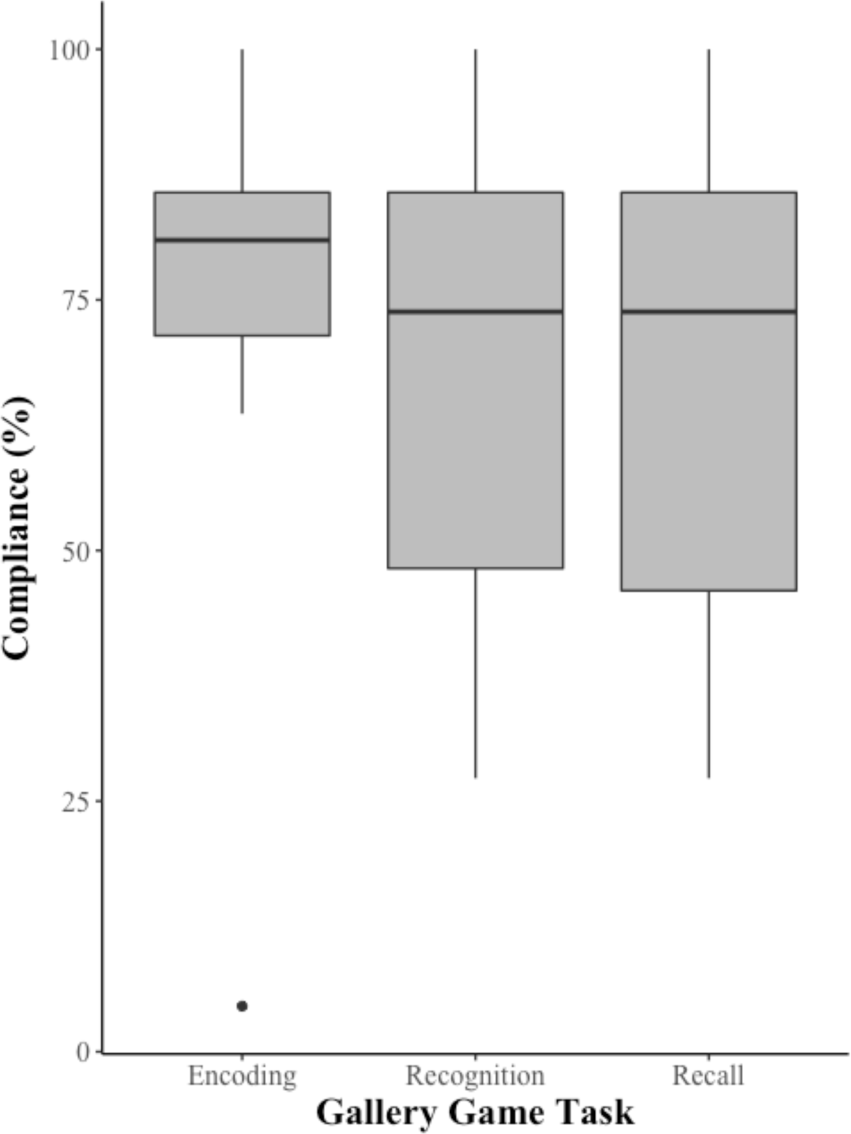
Proportion compliance for Gallery Game tasks (learning, recognition and recall) across the baseline month of assessments.

### Thematic analysis of app experience

Quotations evidencing the extracted themes are found in Table 2.

**Table 2.**
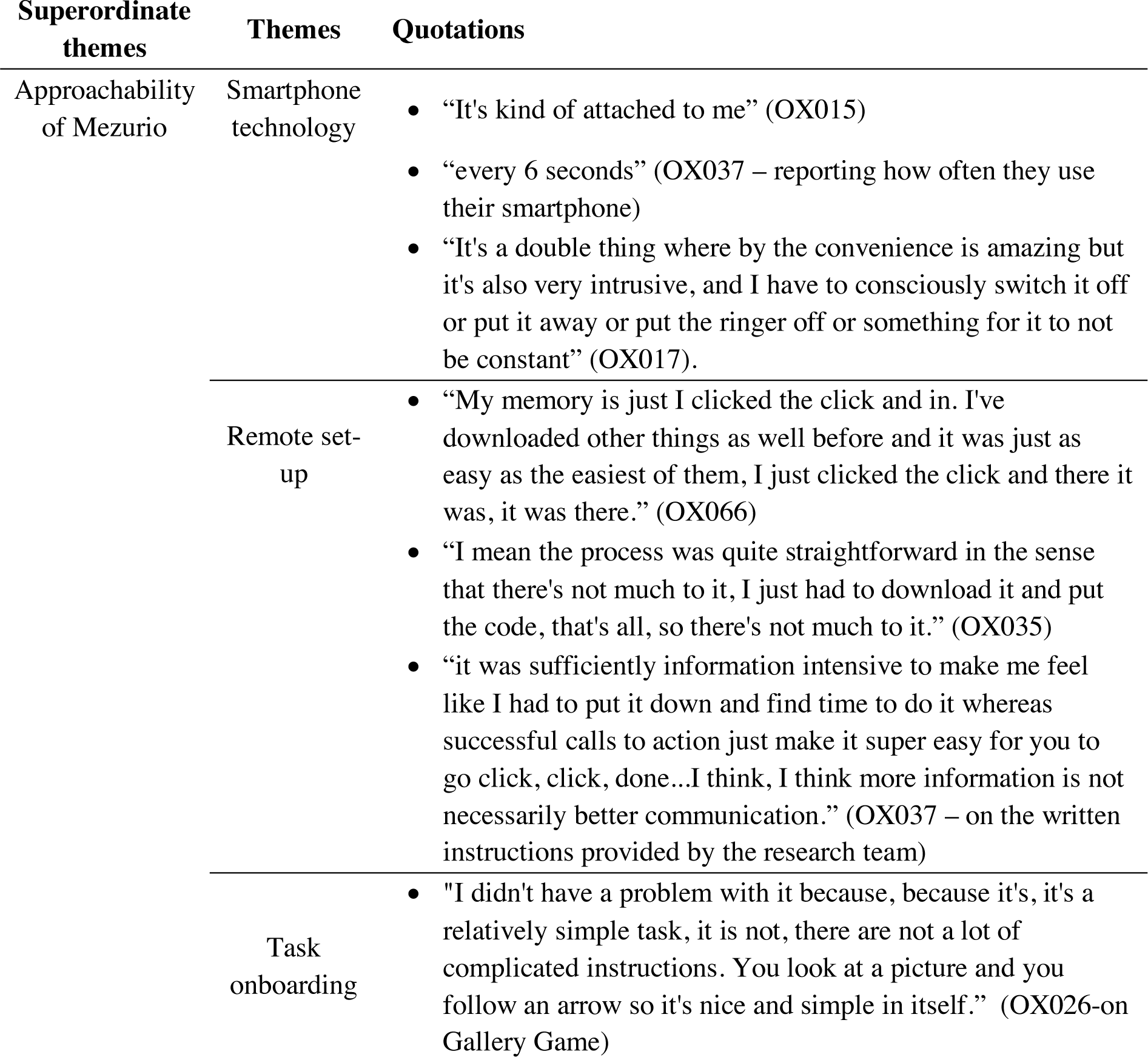
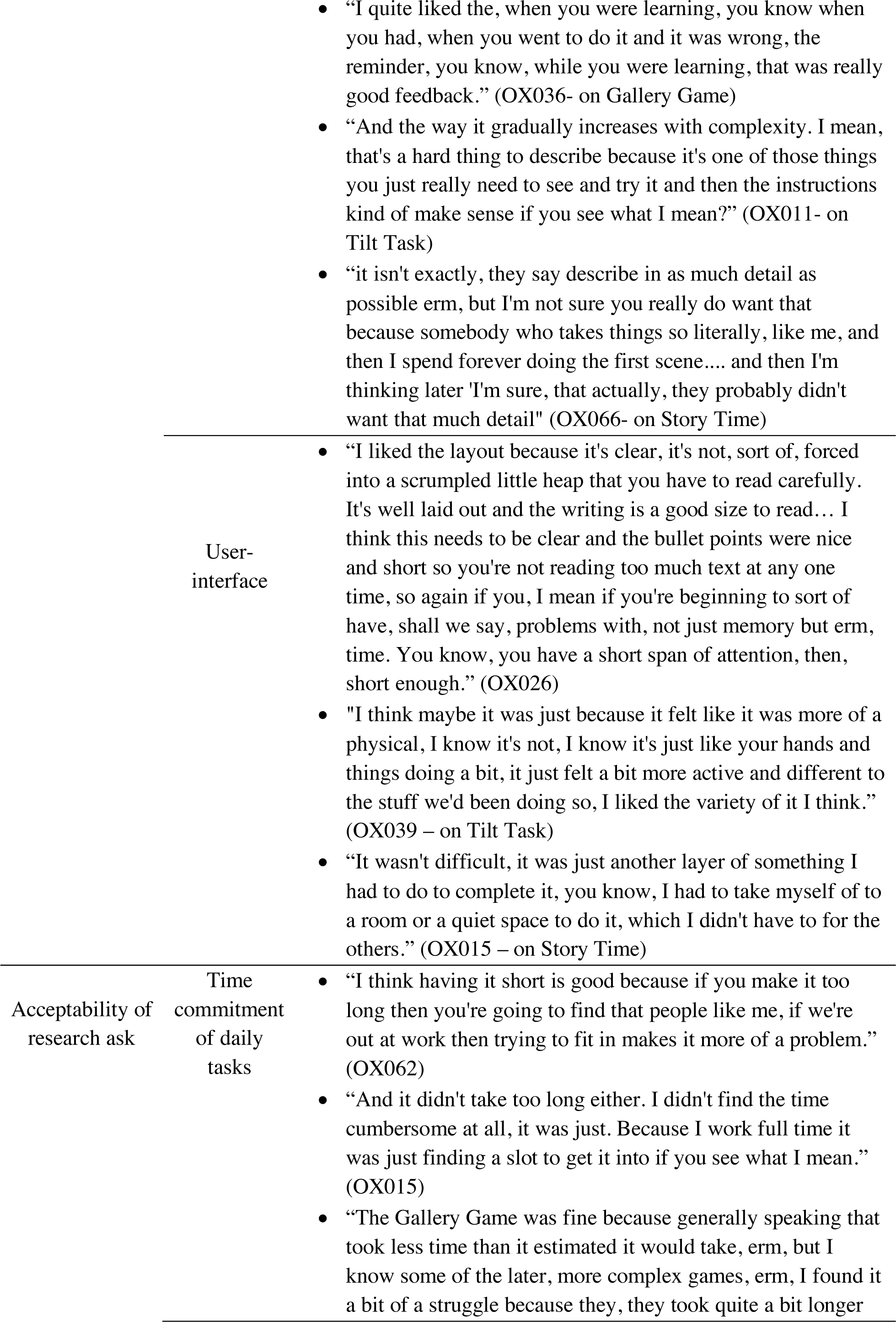
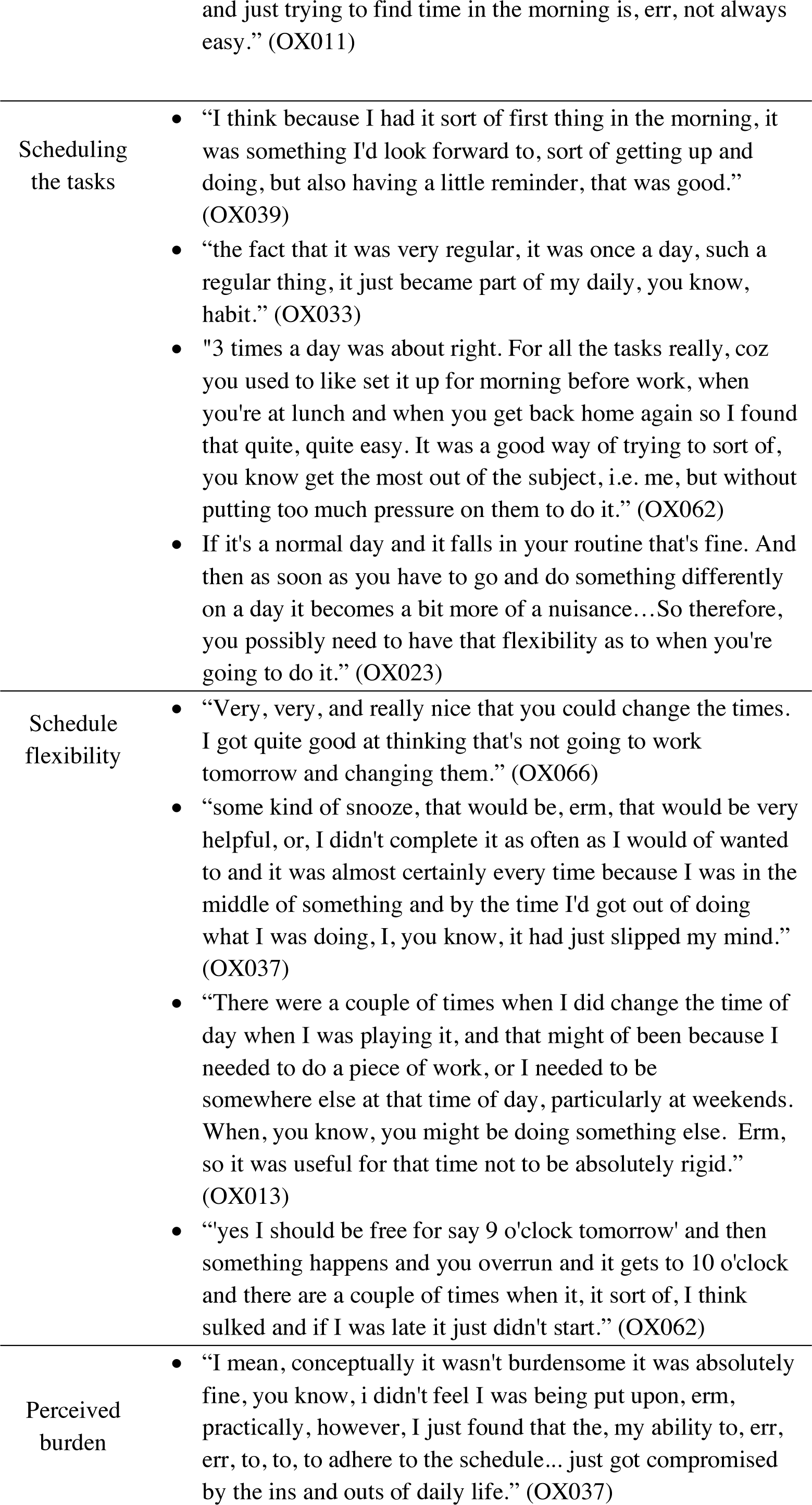
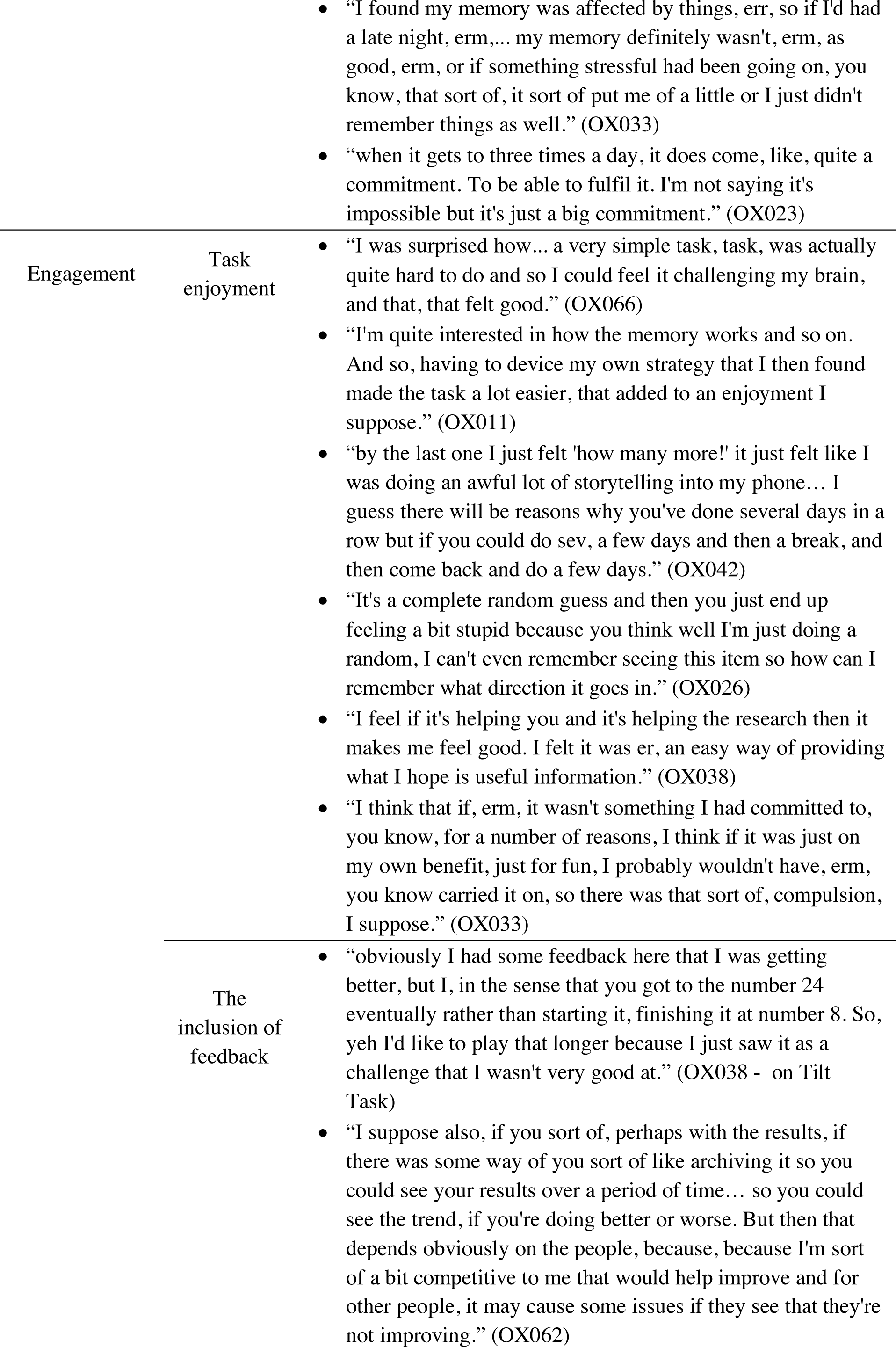
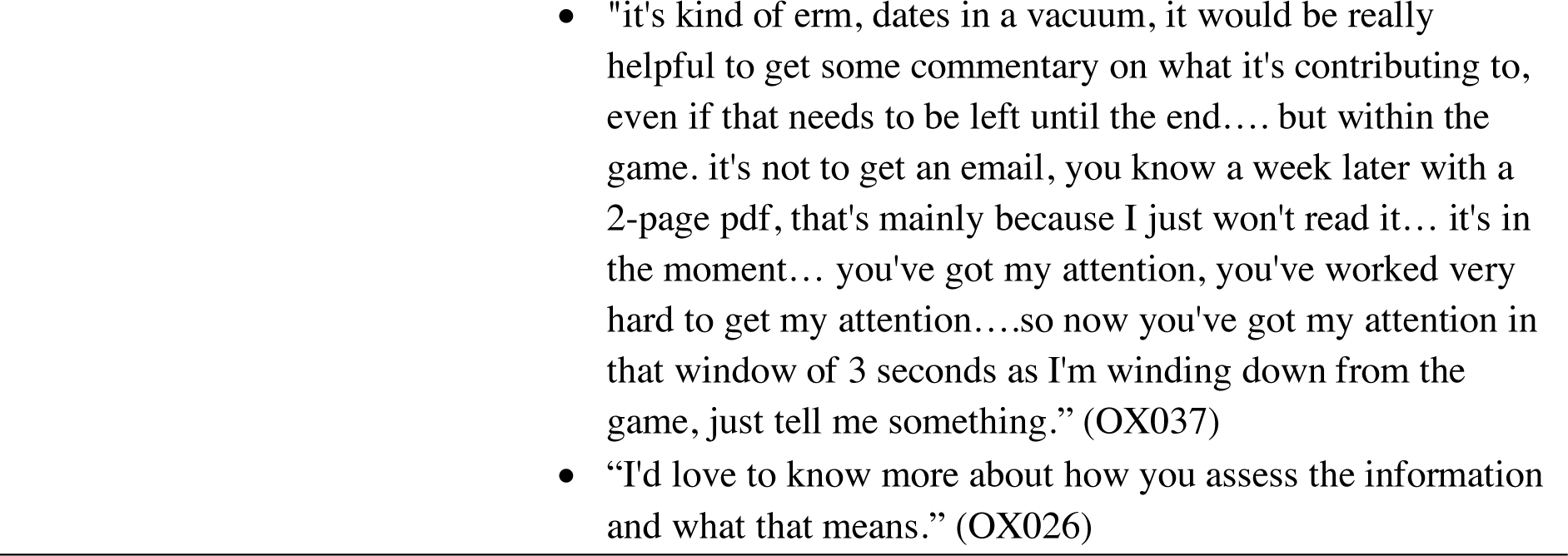
Thematic outcomes of self-reported research experience with the Mezurio app, evidenced by participant quotations.

### Approachability of the Mezurio app

**The approachability of smartphones** was confirmed in the current mid-age sample, with all participants owning a personal device (12 Apple iPhone-users; 8 Android-users) and 90% reporting daily or more frequent usage (the remaining participants stated they used their phone most days). Indeed, 7 participants reported active attempts to decrease smartphone usage in their daily lives supporting the prevalence of these technologies in the target population. Smartphones were used by participants for a wide range of functions, including communication, work, finance, travel and shopping, however, 35% of participants reported viewing their phone as a functional tool rather than a device used for enjoyment. The majority (*n*=15) of this mid-age populations did not use their phones for gaming, with the two reasons reported for this being time commitment (*n*=3) and lack of interest (*n*=3). A small number had previously completed AD-focused SeaHero Quest (*n*=2) (Coutrot et al., 2019) or “brain-training” apps (*n*=2).

**Remote set-up** of the Mezurio app was well accepted, with the clarity of the installation process scored an average of 9.03/10 ±1.52 and 80% of participants reporting written instructions for installation were sufficient; indeed, one participant felt the quantity of written guidance provided by the research team was a barrier to them completing what in reality was a very simple task.

**Task onboarding** for Gallery Game was appropriate for the current sample with instructions scored an average of 9.30/10 ± .10. Participants highlighted a number of features which were effective for remote introduction to Gallery Game, including simple, short written instructions, the inclusion of ‘pop-up’ responsive help within the task, and the provision of a task strategy at the start of the assessment period. Participants were allowed up to 7 days to practice the Gallery Game task; the consensus (*n*=13) was that an initial practice period of 1 or 2 days is sufficient for understanding task demands. For participants completing Tilt Task, again the instructions were judged to be clear, with participants highlighting the gradual increase in task complexity coupled with an opportunity to practice as important factors. There was less consensus around the clarity of task onboarding for Story Time; participants were unclear of how much descriptive detail to provide during narration.

**The User-interface** of Gallery Game and Tilt Task was acceptable, with the manual response mechanism of the Tilt Task evaluated positively by 8 participants. Qualitative feedback suggested the spoken interface of Story Time increased the burden of remote, digital assessment (*n*=7) by limiting situations in which the task could be completed. Across Mezurio, visual clarity of the app was highlighted (*n*=6) as an important feature.

### Acceptability of the research ask

The current approach to remote cognitive assessment was scored 8.37 out of 10 ± 1.34 for participant acceptance. The limited number of participants (*n*=4) completing the Gallery Game only version of Mezurio precludes traditional significance testing, however mean acceptability of this group (*M*=9.00) is comparable to ratings by those completing the extended version of the app (*M*=8.20).

**Time commitment of the daily tasks** was an important feature in the acceptability of the current participation ask, with the duration of tasks (∼ < 5 minutes) being important in allowing the app to work around participants’ schedules. Increased daily participation load in the extended version of Mezurio, characterised by occasional additive scheduled activities (e.g. a Gallery Game and Story Time on the same day) with longer daily durations contributed to an increased sense of burden (*n*=3). The current study asked participants to interact with the app every day for up to 36 days; this commitment was not a significant problem for the current research group with limited (*n*=3) discussion of this as an issue.

**Scheduling the tasks** via phone-based notifications benefitted study compliance (*n*=30), with many participants noting the regularity of app timings helped them complete their daily tasks. Specifically, the inclusion of short, cognitive assessments within participants’ daily routine was highlighted as important, with poor compliance generally accounted for by changes to individual’s regular patterns of activity e.g. weekends, trips (*n*=3). Participants using the extended version of Mezurio were prompted to complete Tilt Task three times a day, with the alignment of task notifications to daily routine (e.g. breakfast, lunch, dinner) reported to be increasingly important for higher-frequency assessment. Three participants reported finding the three times a day too much to fit into their daily routine.

**Flexibility of the study schedule** was emphasised as a requisite for high-frequency, remote assessment. Within the app participants had the option of changing the time of their next prompt, with 7 participants identifying this feature as a strength. Participants (*n*=5) suggested Mezurio could be improved in future through the inclusion of a ‘snooze’ function for notifications, allowing participants to set a second reminder for later in the day. Actively maintaining the intention to complete Mezurio after non-compliance with the initial prompt was associated with increased subjective burden (*n*=2). At present, participants are able to re-enter the Mezurio app and complete their daily task within a certain expiry window (once daily tasks: 16 hours, thrice daily tasks: 2 hours). This feature benefitted research acceptability, however, there was a wish for still greater flexibility. A further suggestion to improve Mezurio is the ability for participants to monitor task expiry time (*n*=3).

**Perceived burden** of the research ask was to a large extent dependent on additional stressors to a participants’ time external to the app, with daily life stressors associated with subjective reports of poorer cognitive performance on that day. Asking participants to interact with the app three times a day was considered burdensome by some participants (*n*=3).

### Engagement

**Task enjoyment** was scored (/10) as follows: Gallery Game *M*=7.26 ± 2.02, *n*=19; Tilt Task *M*=6.44 ± 2.62, *n*=12; Story Time *M*=7.18±1.49, *n*=14. The cognitive demand of included assessments was cited as a primary source of participant enjoyment (*n*=9), with participants reflecting on the satisfaction of challenging themselves in comparison to their previous performance and the development of personal strategies to aid performance (*n*=3). In contrast, barriers to research enjoyment include worry about their own performance (*n*=6), frustration at difficult tasks (*n*=4), and limited variation in day-to-day task demand (*n*=2). Enjoyment was not considered an important factor for compliance by a small number of participants describing the tasks as functional as opposed to fun. In relation to this, a main motivator for engagement was commitment to the research aims rather than personal pleasure.

**The inclusion of feedback** was identified by participants as the foremost way to promote participant engagement with remote cognitive assessment. Although limited explicit feedback on performance outcomes is provided in the current version of Mezurio, participants had an intuitive sense of their own performance (*n*=7). Participants highlighted a need for explicit feedback on how their performance changes across the course of the research, with feedback on performance in comparison to peers identified as a potential factor to increase study participation. In addition, greater dissemination of the research background, objective and methods within the app was highlighted as a potentially to promote further engagement.

## Discussion

The current study tested the feasibility of digital cognitive assessment, deployed through the Mezurio smartphone app, in a mid-age group relevant to the detection of early preclinical AD (Finch, 2009; Irwin, Sexton, Daniel, Lawlor, & Naci, 2018). Participants were prompted to complete a prolonged schedule of daily assessments (≤ 36 days); qualitative evaluation of Mezurio’s user interface and task design, alongside study compliance and attrition, were used to explore if smartphone-centred tools can substantially extend the breadth of cognitive assessment in which participants will engage. Excluding one participant who subsequently withdrew due to the time commitment, compliance with the schedule of Gallery Game learning tasks averaged 80%, with 88% of participants still active at the end of the assessment period (≤26 days), confirming the feasibility of frequent, long-term cognitive assessment in a digital environment. Furthermore, participant feedback supported the acceptability of Mezurio’s approach to digital assessment, with an intuitive user-interface, flexible scheduling around personalised prompts, and engagement within the tasks themselves identified as important factors contributing to a positive research experience.

Importantly, the present study demonstrated participant engagement with daily cognitive assessments across a significantly longer study duration than the previously reported seven-day window (Allard et al., 2014; Lange & Süß, 2014; Schweitzer et al., 2017). This provides evidence that smartphone technologies enable far greater sampling of cognition than plausible with in-person tests. Consistent with the high-levels of participation reported in older adults asked to complete frequent mobile behavioural assessments (Allard et al., 2014; Lange & Süß, 2014; Schweitzer et al., 2017), high compliance in this mid-age group confirmed the feasibility of the current approach to smartphone-based cognitive testing. Furthermore, limited attrition supports the potential utility of smartphone-based tools for longer-term monitoring. Participation ‘drop-out’ is reported in digital research spanning multiple months (Jongstra et al., 2017; Valdes, Sadeq, Bush, Morgan, & Andel, 2016), with cited reasons for withdrawing from the research including technical problems, time commitment and loss of interest in repetitive tasks (Valdes et al., 2016). Whilst study reuptake at months 6 and 12 for this initial pilot of Mezurio remains to be established, progression of strategies to sustain participant engagement is important for the quality of collected data.

A major benefit of smartphone assessment is the ability for participants to independently contribute to research from a home environment, hence it is critical to establish the remote usability of such measures prior to implementing them at scale. Mid-age participants in the present study positively evaluated the clarity of set-up and task instructions for Mezurio, with concise written instructions, an opportunity to practice and responsive inbuilt help identified as design strengths - developed through iterative patient and public involvement. All participants were frequent smartphone users, however, and although the adoption of smartphone technology is increasing in older age-groups (49% of UK adults aged 55-64 years and 17% of over 65s reported owning a device in 2015 (Ofcom, 2015)), generalisability of app usability remains to be established. Subjective feedback from participants, including those with no prior experience of smartphones, on a smartphone app for monitoring the symptoms of chronic pain identified inexperience with technology and the need for technical support as potential issues (Reade et al., 2017). Future work will further establish the feasibility of Mezurio in a wider general population.

Scheduling, both the length of individual assessments and participant autonomy in the timing of smartphone-based notifications, was critical for the acceptability of reseach ask (Jenkins et al., 2016; Koo & Vizer, 2019; Van Middelaar et al., 2018). Each task within Mezurio is designed to take less than five minutes to complete; whilst acceptability of the current participation ask was high (8.37/10), time commitment was reported as a primary reason for non-compliance emphasising the value of a ‘little and often’ approach for repeat, mobile assessment (Allard et al., 2014; Lange & Süß, 2014; Schweitzer et al., 2017). Allowing participants to personalise phone-based notifications to suit their daily routine and flexibility to delay responding to scheduled assessments were identified as important factors in limiting research burden.

The mental challenge of daily activities was identified as an important factor for participant engagement with repeat Mezurio assessments, with the suite of tasks within this app intended for use by adults with no clinical diagnosis of cognitive impairment in contrast to a more traditional approach to neuropsychological assessment. In a limited number of participants, however, subjective performance was associated with anxiety, relevant for future modification of these tasks for use in individuals with mild cognitive impairment. At present, Mezurio provides limited explicit feedback on task performance, with participants suggesting future personalisation of this aspect of the app would benefit sustained participant engagement, consistent with previous focus groups (Jenkins et al., 2016; Van Middelaar et al., 2018). In accordance with Mezurio’s intended screening utility for preclinical dementia, careful consideration of how to ethically communicate performance outcomes to participants is needed. Importantly, present feedback suggested short communications within the app intended to inform and promote interest in the research area may likewise contribute to long-term engagement.

One limitation of the current study is that the Tilt task and Story Time were included as an optional, later amendment, meaning participants did not have a uniform experience of using the app. In addition, reasons for participants declining to take part in both the extended version of Mezurio at 6-month follow-up, and in this mobile technology sub-study of PREVENT more broadly was not recorded which would have provided additional insight on the approachability of smartphone cognitive assessment. When drawing on these results it is worth noting the current sample was recruited from an existing, more intensive prospective cohort, indicative of a high commitment to dementia research which was mirrored in qualitative feedback. Given the promise of using digital biomarkers to screen risk remotely in advance of clinical consultation, it is important to extend these results to demonstrate feasibility in a general population. The GameChanger study (https://joingamechanger.org) has been launched directly address this need, with over 16,000 participants from the general UK population completing remore, frequent cognitive assessments with the Mezurio app to date. It is also notable that the recruited sample consisted of cognitively healthy individuals and thus the acceptability/feasibility of using the app in impaired populations remains to be studied. Conclusions from the present research, alongside further input from patient and public involvement sessions, has been used to strengthen the research design implemented in the Mezurio smartphone app.

## Conclusions

This research confirmed the feasibility of the Mezurio smartphone app for extensive cognitive profiling; evidencing high compliance to a substantially longer schedule of daily interactions than previously explored. The scheduling of smartphone interactions, clarity of user experience and task design were critical for reported engagement with Mezurio’s research design, with the qualitative feedback presented here providing important direction for implementing digital tools in future health research. Following this initial demonstration of the viability of remote Mezurio assessment as a complementary method to in-clinic assessment, ongoing work seeks to replicate this feasibility in a wider population, plus adults with some degree of cognitive impairment. In addition, the scientific utility of these novel cognitive tasks in comparison to traditional markers of preclinical dementia (brain-based biomarkers, neuropsychological outcomes, prospective decline) is in progress. The present conclusions, however, are an important first step in justifying a participant-orientated, mobile design to progress efficient, early screening of cognitive decline.

## Data Availability

The data contained within this article is not publicly available.

## Acknowledgements

Development of the Mezurio app was supported by funding from the Robertson Foundation, NIHR Oxford Health Biomedical Research Centre, Eli-Lilly and F. Hoffmann-La Roche Ltd. Additional support for this study is provided by the Oxford Clinical Academic Graduate School Clinical Lecturer Support Scheme, the Academy of Medical Sciences Clinical Lecturer Starter Grant (SGL016\1079) and the Medical Research Council Deep and Frequent Phenotyping study grant (MR/N029941/1). The PREVENT research programme was developed with a grant from Alzheimer’s Society.

## Appendix – Interview Schedule

1. Was this your first time using a smartphone?
  a. If yes:
    i. Did you enjoy using the smartphone?
    ii. Could you describe any problems you had using the smartphone?
  b. If no:
    ▪ If you own a smartphone, is it Android or Apple?
    ▪ How regularly do you use a smartphone?
    ▪ What sort of things do you use your smartphone for?
    ▪ Do you have experience playing games on a smartphone?
    ▪ If the participant says they play games – what sort of games, how often and why? If not, why not?
    ▪ How enjoyable do you find using your smartphone?
  **Downloading, installing and setting up the app:**
2. How do you rate the app download process out of 10, with 10 being very straightforward and 1 being very complicated?
  - Why? e.g. was there sufficient information from the research team, any problems downloading and installing the app.
  **Instructions and daily notifications:**
3. How clear did you find the instructions for completing the Gallery Game task on a scale of 1-10, with 10 being very clear and 1 being not clear at all?
  - Why? e.g. what was unclear, did you have any problems understanding the instructions?
4. The app sent you a notification each day to remind you to play the game, were these helpful in completing your task?
  - Is there anything else we could do to help you engage with the app every day?
  **Gallery Game**
5. The app asked you to complete a task, for a few minutes, every day for about a month. How acceptable or burdensome was this on a scale of 1-10, where 10 is very acceptable, and 1 is very burdensome?
  - Why?
6. Were there any occasions when you weren’t able to complete the task?
  - Why?
7. How enjoyable did you find doing the memory task on a scale of 1-10, with 10 being very enjoyable and 1 being not at all enjoyable?
  - Why?
8. How many practice days did you need before you felt confident completing the task?
9. Did you use a strategy to help you remember photos during the memory task?
  a. If so, can you describe it?
10. How easy or difficult did you find the completing the memory task on a scale of 1-10, with 10 being very easy and 1 being very difficult?
  - Why? e.g. which aspects did you find difficult?
  **For people who consented to complete the extended version of Mezurio** **The Tilt Task**
11. How enjoyable did you find doing the Tilt task on a scale of 1-10, with 10 being very enjoyable and 1 being not at all enjoyable?
  - Why?
12. Did you feel the practice days were sufficient for you to understand the task?
13. How easy or difficult did you find the completing the Tilt task on a scale of 1-10, with 10 being very easy and 1 being very difficult?
  - Why? e.g. which aspects did you find difficult?
  - Aspects of the task you would improve if they say it was difficult for technical/app-based reasons rather than the cognitive demand of the task itself?
  **Story time**
14. How enjoyable did you find doing the Story tie on a scale of 1-10, with 10 being very enjoyable and 1 being not at all enjoyable?
  - Why?
  Suggestions for improvement if found the task not so enjoyable
15. How easy or difficult did you find the completing the memory part of Story time on a scale of 1-10, with 10 being very easy and 1 being very difficult?
  - Why? e.g. which aspects did you find difficult?
  **General Questions:**
16. How would you rate the experience of using the app out of 10, with 10 being a very positive experience and 1 being a very negative experience?
  - Why?
17. Can you describe any problems or issues you had with the app?
18. What aspects of the app did you like?/dislike?
19. Is there anything we could do to improve your experience of using the app? (Including downloading the app, instructions and the design of the tasks)

